# Neonatal hypothermia and adherence to World Health Organisation thermal care guidelines among newborns at Moi Teaching and Referral Hospital, Kenya

**DOI:** 10.1101/2020.06.03.20121053

**Authors:** Winstone Mokaya Nyandiko, Paul Kiptoon, Florence Ajaya Lubuya

## Abstract

**Background:** Neonatal hypothermia is a great concern with near epidemic levels globally. The prevalence in Kenya is as high as 87%. Local data on the associated factors including adherence to warm chain guidelines as recommended by the World Health Organisation (WHO) is limited.

**Objective:** To determine the prevalence of hypothermia and level of adherence to the WHO thermal care guidelines among newborns admitted at Moi Teaching and Referral Hospital (MTRH).

**Methods:** This descriptive cross-sectional study was carried out among neonates admitted at the MTRH newborn unit. Systematic sampling was used to recruit 372 eligible participants. Axillary thermometry, interview of respective mothers and observation of thermal care practices was done. Means and medians described continuous variables while frequencies with corresponding percentages summarized categorical variables. Associations between various variables and neonatal hypothermia were computed using the Pearson chi-square test. Relative Risks and Odds Ratios were assessed between predictor and outcome variables. Independence among significant variables was determined through the logistic regression model at 5% prediction level.

**Results:** Among the 372 participants, 64.5% (n=240) were born at MTRH, 47.6% (177) were preterm while 53.2% (198) had birth weights below 2500 grams. Admission hypothermia was noted among 73.7% (n= 274) while 13% (49) died on day one of admission. Only 7.8 % (29) newborns accessed optimal thermal care. Prematurity (RR=1.62, 95% CI: 1.43-1.84), day one mortality (RR=17.7, 95% CI: 2.40, 122.71) and adherence to the warm chain (p<0.001) was significantly associated with admission hypothermia. Inappropriate thermal resuscitation appliance (RR=1.50, 95% CI: 1.34-1.67) inappropriate clothing (RR = 1.78, 95% CI: 1.54 - 2.05) and late breastfeeding (RR = 2.01, 95% CI: 1.39-2.89) significantly increased the risk of hypothermia. Non hypothermic newborns had twenty-fold increased odds of survival (AOR=20.91, 95% CI: 2.15-153.62).

**Conclusion:** Three out four neonates at the MTRH newborn unit had hypothermia at admission. Hypothermia was significantly associated with prematurity, adherence to warm chain and day one mortality. There was notably low adherence to the warm chain.

**Recommendation:** Strategies to optimize adherence to the warm chain at MTRH with emphasis on ^1^thermal care of the preterm neonate should be instituted.

## INTRODUCTION

Neonatal hypothermia is defined by the World Health Organisation (WHO) as an axillary temperature below 36.5°C (97.7°F) among newborns aged below 28 days[1]. It is stratified as either mild (36°C-36.4°C), moderate (32°C-35.9°C) and severe hypothermia (<32°C) with the severity scale carrying prognostic implications[2]. Newborns lose heat through conduction, radiation, convection or evaporation[3]. Neonatal hypothermia has been associated with a number of risks including inherent factors such as physiological and behavioural characteristics or external factors such as the environmental conditions[4]. In Spain, hypothermia was associated with VLBW among infants[5]. Similar results was seen earlier in a study in NICUs in Iran[6]. Previous evidence from sub-Saharan Africa indicate sub-optimal thermal care practices, inadequate thermal education among providers and community level determinants of neonatal hypothermia[7,8].

To reduce hypothermia associated neonatal deaths, the WHO proposed a ten-step ‘warm chain’ guideline that includes:[9] Warm delivery rooms with temperatures between 25°C to 28°C at the birthplace; Immediate drying before the delivery of the placenta using pre-warmed towels; Skin to skin contact (SSC) for mother and baby that is among principles of kangaroo mother care (KMC); Early breastfeeding (within one hour) or at least on the first day of life; Delayed weighing and bathing for at least 6 hours and 24 hours respectively; Appropriate clothing and bedding (*with at least 3 layers of dry and absorbent material*); Rooming-In by keeping mother and baby together; Warm transport-use of warm wrap, external heat source and SSC; Warm resuscitation (*using appropriate appliances during resuscitation*) and Continued thermal care training for parents, caretakers and health workers.

## MATERIALS AND METHODS

This was a cross-sectional study done at the Relay Mother and Baby Hospital’s (RMBH) newborn unit (NBU) of Moi Teaching and Referral Hospital (MTRH) in Eldoret-Kenya between July and December 2016. The study systematically recruited 372 neonates admitted to the NBU within the first hour of admission on their first day of life.

Low reading axillary thermometers (32°C to 42°C) and ambient air thermometers were sourced from a supplier with prior accreditation by the hospital. Eligible neonates were enrolled after completion of the admission procedure. After administering an informed consent to the mothers, neonatal axillary temperatures were taken and recorded in a pretested questionnaire where maternal sociodemographic, clinical and thermal care related details were collected. A checklist of the available equipment, ambient temperature and observed aspects of thermal care practice was filled. Serial temperatures were taken at the 1^st^, 3^rd^, 6^th^, 12^th^ and 24^th^ hour or at the point of last contact with the newborn on day one of admission. Critical temperature values were reported to the care team and the medical records were reviewed and updated appropriately.

Descriptive statistics techniques including measures of central tendency (means, medians, frequencies and corresponding proportions) were used to describe the study participants. Inferential statistics involving Pearson’s chi-square tests of association, Risk and Odds Ratio were used to draw associations among the predictor and outcome variables. Ethical approval to carry out the study was obtained from the Institutional Research and Ethics Committee (IREC) of Moi University and the MTRH management.

## RESULTS

### Neonatal characteristics

A total of 372 neonates with an average gestational age of 35.4 weeks (±3.9) were enrolled into the study. Among them, 57.3% (n=213) were males while 47.6 % (n=177) were preterm. Day one mortality was observed among 13.2% (n=49) of the participants (Table 1).

**Table 1:** Neonatal demographic and clinical profile

|  | n(%) / Mean (SD) or Median (IQR) |
| --- | --- |
| **Age (Minutes), Median (IQR)** | 83 (34, 246) |
| Range (Min. - Max.) | 6 – 1410 |
| Gestational age (weeks) Mean (SD) | 35.4(3.9) |
| Range (Min-max) | 26-41 |
| **Gender (n%)** |  |
| Male | 213 (57.3%) |
| Female | 159 (42.7%) |
| **Place of birth (n%)** |  |
| MTRH | 240 (64.5%) |
| Non MTRH | 132 (35.5%) |
| **Mode of Delivery** |  |
| Operative | 100 (26.9%) |
| Non-operative | 272(73.1%) |
| **Maturity by NBMS** |  |
| Term | 195 (52.4%) |
| Preterm | 177 (47.6%) |
| **Maturity by birth weight** |  |
| <2500g | 198 (53.2%) |
| $\geq 2500$g | 174 (46.8%) |
| **Diagnosis** |  |
| Prematurity | 128 (34.4%) |
| Birth asphyxia | 135 (36.3%) |
| Surgical/congenital disease | 43 (11.6%) |
| Macrosomia | 6 (1.6%) |
| Presumed sepsis | 60(16.2%) |
| **Day one outcomes** |  |
| Admission hypothermia | 274 (73.7%) |
| Hypothermia recurrence | 254 (68.3%) |
| Day one mortality | 49(13.2%) |
| **Proportion of Neonatal Hypothermia** |  |

### Proportion of Neonatal Hypothermia

Nearly three quarters 73.7% (n= 274) of the neonates were hypothermic while only 7.8% (n=29) accessed optimal thermal care within the 1^st^ hour of admission. More than half (68%; n=252) of the neonates had recurrent episodes of hypothermia on the first day of admission (Table 1). Among the neonates enrolled, majority of them had moderate hypothermia (46%) with one-tenth recording severe hypothermia (Figure 1).

**Figure 1:**
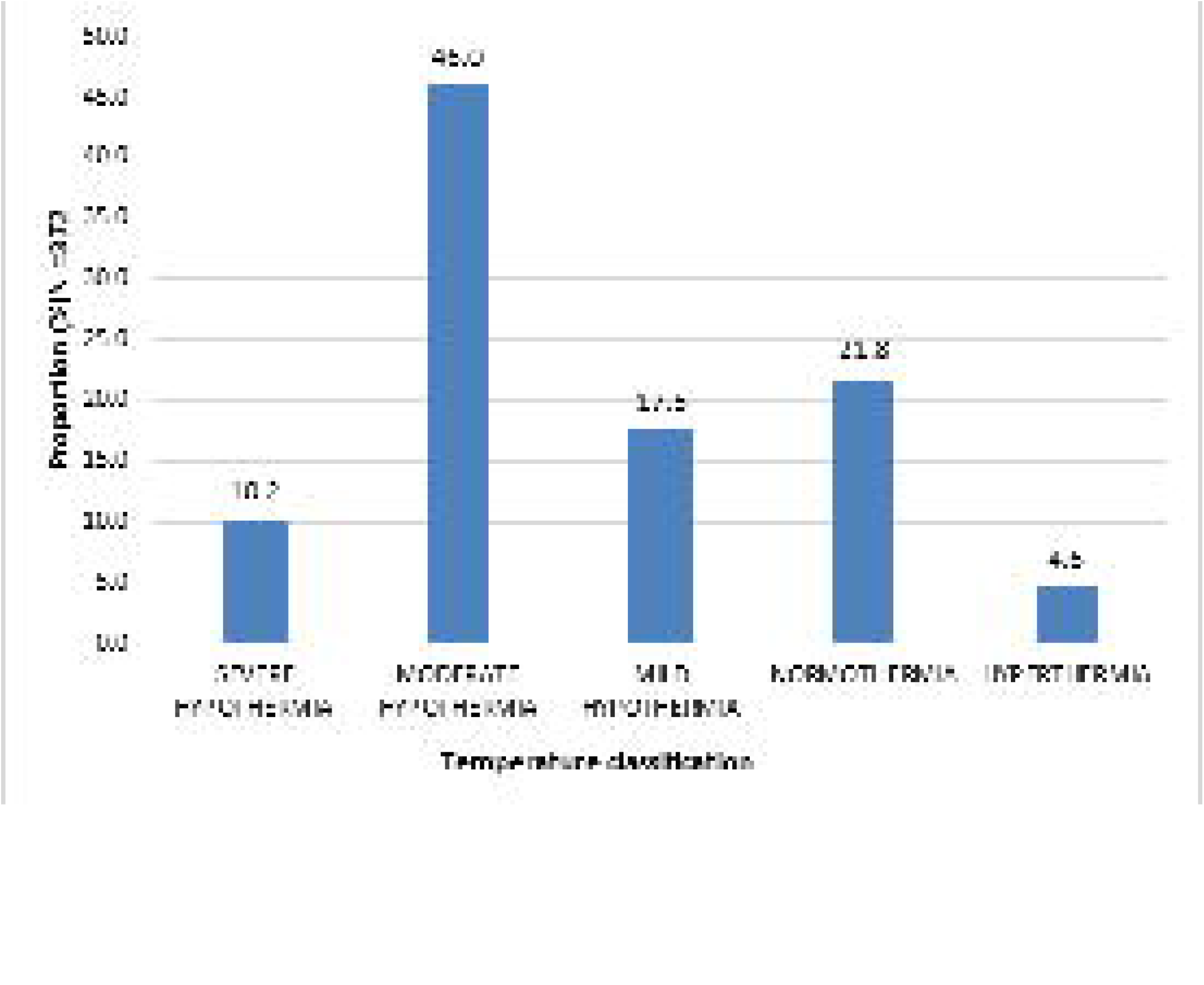
Spectrum of neonatal temperatures at the 1st hour of admission.

Nevertheless, a steady decline in the prevalence of hypothermia was noted during the initial 24 hours of admission. Serial thermometry revealed hypothermia proportions of 55.6% (n=207), 44.9% (n=167), 39.8% (n=148), 34.9% (n=130) and 23.4% (n=87) at the 3^rd^, 6^th^, 12^th^, 18^th^ and 24^th^ hours respectively (Figure 2). Less than one-fifth of the neonates were not assessed between the 6^th^ and 24^th^ hour because they were out of the newborn unit due to specialized investigations or they had been released back to their mothers in the postnatal ward.

**Figure 2:**
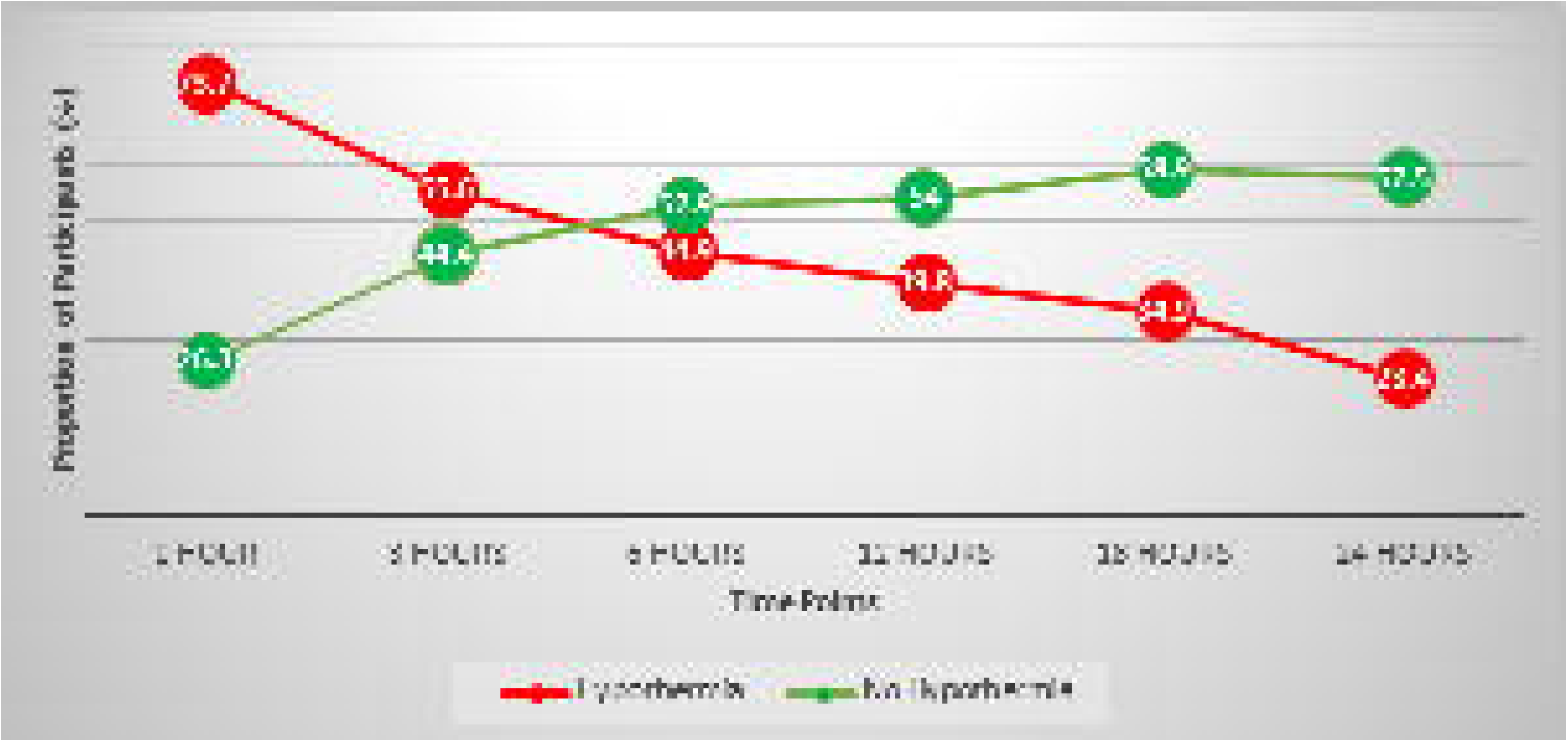
Neonatal hypothermia trends over the initial 24 hours of admission.

### Adherence to individual warm chain steps by the 1^st^ hour of admission

Provision of warm delivery rooms and newborn units at MTRH was assessed over the study period with median ambient temperatures of 24.53°C (IQR: 22°C, 27°C) {n=38}, 20.13°C (19°C, 22°C) {n=32} and 25.4°C (24°C, 28°C) {n=62} recorded at the labor rooms, operating theatres and newborn units respectively. Almost half (43.8%; n=163) of the mothers delivered in warm rooms irrespective of the place of birth, while majority of the neonates (81.7%; n=304) were wiped and wrapped immediately. Warm transport was observed among 243 (65.3%) neonates at admission with 82% (n=305) of them dressed in three layers of dry, warm and absorbent clothing. However, there was notable absence of caps and stockings predominantly among the preterm babies. This lowered the aggregate score for appropriate clothing to 48.9% (n=182) as only 53 % (n=197) neonates wore a cap and stockings. Baths were delayed among 73.7% (n=274), while the provision of appropriate thermal resuscitation appliances was noted among 63.7% (n=237). About two fifths (40.3%; n=150) of the mothers had not received any thermal care education. Rooming in (4.6%; n=17), skin to skin contact (9.4%; n=35) and early breastfeeding (12.5%; n=46) were among the least adhered to steps besides the near universal immediate weighing (94.1%; n= 350) as shown on table 2.

**Table 2:** Adherence to WHO thermal care guidelines by the 1st hour of admission, (N=372)

| **Warm chain step** | **YES (%)** | **NO (%)** |
| --- | --- | --- |
| Warm delivery rooms/Newborn Units<br>*Mean ambient temperatures at MTRH (IQR)* | 163(43.8) | 209(56.2) |
| ▪ Labour wards (n=38): 24.53°C (22- 27)<br>▪ Obstetric theatres (n=32): 20.13°C (19-22)<br>▪ Newborn unit (n=62): 25.4°C (24°C-28°C) |  |  |
| Immediate wiping and wrapping | 304(81.7) | 68(18.3) |
| Warm transport | 243(65.3) | 129(34.7) |
| Delayed weighing | 22(5.9) | 350(94.1) |
| **Delayed Bathing** | **274(73.7)** | **98(26.3)** |
| **Appropriate clothing** | **182(48.9)** | **190(51.1)** |
| At least 3 layers | 303(81.5) | 69(18.5) |
| Dry, warm and absorbent | 305(82.0) | 67(18) |
| Cap /stockings | 197(53.0) | 175(47.0) |
| **At least 1/3 warm chain steps under KMC** | **65(17.5)** | **307(82.5)** |
| Early breastfeeding | 46(12.5) | 326(87.5) |
| Skin to skin care | 35(9.4) | 337(90.6) |
| Rooming-in | 17(4.6) | 355(95.4) |
| **Appropriate thermal resuscitation appliance** | **237(63.7)** | **135(36.3)** |
| **Continued thermal care education** | 222(59.7) | 150(40.3) |

### Cumulative adherence to warm chain steps feasible post admission

About a half (51.6%; n=192) of the neonates had two to three steps optimally adhered to. Only (5.1% n=19) had optimum adherence to at least five of the post-admission steps (Figure 3).

**Figure 3:**
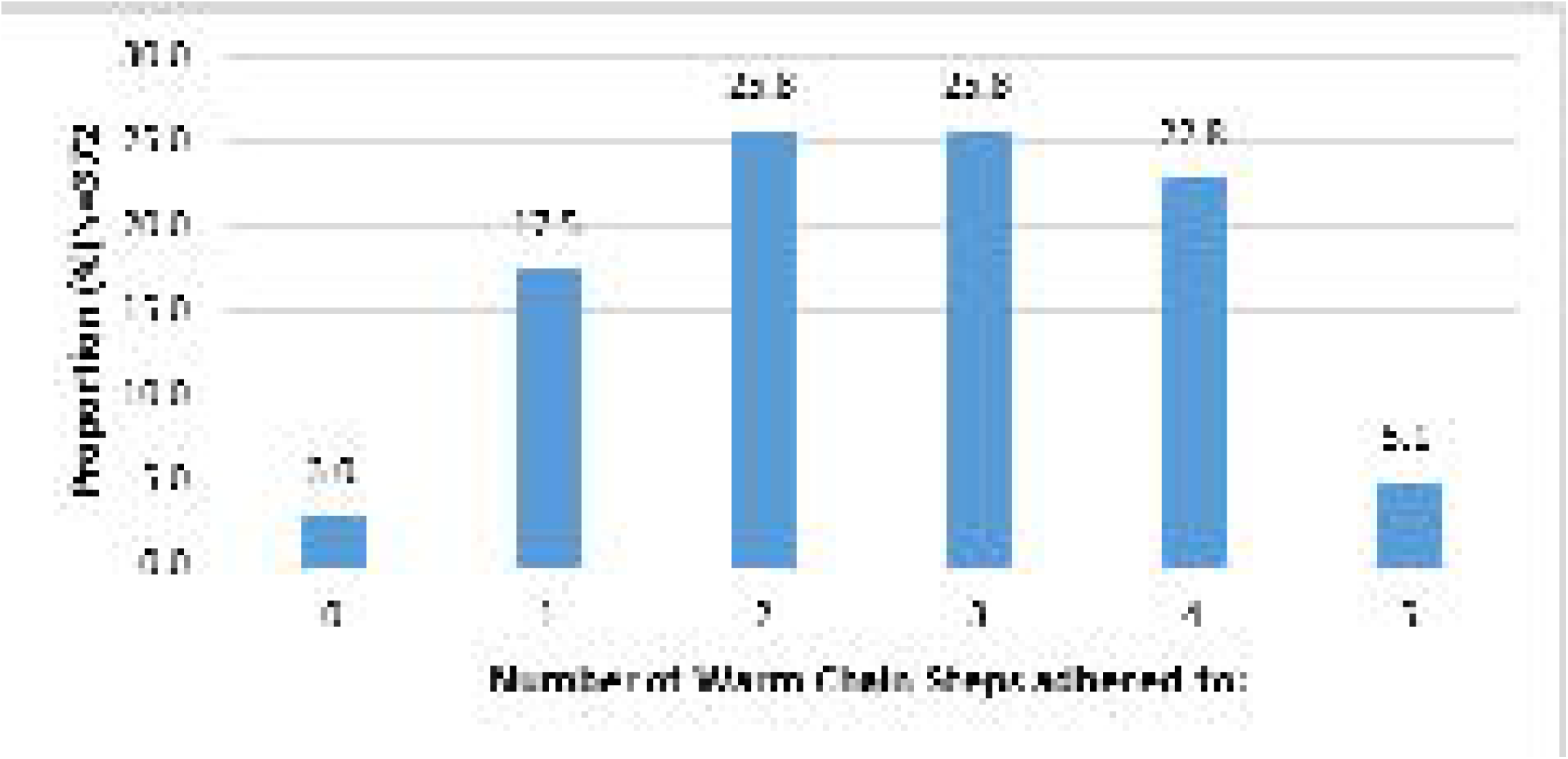
Cumulative adherence to warm chain steps feasible post admission.

### Optimal adherence’ to the warm chain by the 1^st^ hour

There was optimal adherence to the warm chain steps only among (7.8%; n=29) of the participants.

### Demographic and clinical factors associated with admission hypothermia

Birthweight below 2500grams (RR=1.58; 95% CI: 1.37, 1.82) and gestational age below 37weeks (RR=1.62; 95% CI:1.43, 1.84) increased the risk of hypothermia at admission by 58% and 62% respectively. The risk (RR = 1.14; 95% CI: 1.01-1.28) conferred by lack of continued thermal education among the mothers was statistically significant (p-value = 0.041) as shown on table 3.

**Table 3:** Demographic and clinical factors associated with admission hypothermia among newborns admitted at MTRH, Kenya, 2016 [N_D_=_D_372].

| Variables | Admission hypothermia | | P value<br>Critical<br>$P \leq 0.05$ | Relative risk (RR)<br>(95% CI) |
| --- | --- | --- | --- | --- |
|  | YES: n (%) | NO: n (%) |  |  |
| **Place of birth:** Non-MTRH<br>MTRH | 94 (71.2)<br>180 (75) | 38 (28.8)<br>60 (25) | 0.427 | 0.95(0.83-1.08) |
| **Mode of delivery:** Operative<br>Non operative | 75 (75)<br>199(73.2) | 25 (25)<br>73(26.8) | 0.721 | 1.07(0.73-1.6) |
| **Maturity by NBMS:** Preterm<br>Term | 161(91.0)<br>113(57.9) | 16(9.0)<br>82(42.1) | <0.001 | 1.62(1.43-1.84) |
| **Maturity-weight:**<2500grams<br>≥2500grams | 176(88.9)<br>98 (56.3) | 22(11.1)<br>76(47.7) | <0.001 | 1.58(1.37-1.82) |
| **Maternal parity:** Primiparous<br>Multiparous | 141(76.6)<br>133(70.7) | 43(23.4)<br>55(29.3) | 0.198 | 1.08(0.96-1.22) |
| **Thermal Education:** NO<br>YES | 119(79.3)<br>155(69.8) | 31(20.7)<br>67(30.2) | 0.041 | 1.14 (1.01-1.28) |

### Association between adherence to the warm chain and admission hypothermia

Inappropriate appliances (RR = 1.50; 95% CI: 1.34, 1.67) or clothing (RR=1.78; 95% CI: 1.54, 2.05) and sub-optimal adherence to any of the three KMC steps (RR= 1.79; 95% CI: 1.36-2.36) increased admission hypothermia risk by 50%, 78% and 79% respectively. Early bathing (RR =1.05; 95% CI: 0.92-1.20) was not statistically associated (p= 0.452) with admission hypothermia (Table 4).

**Table 4:**
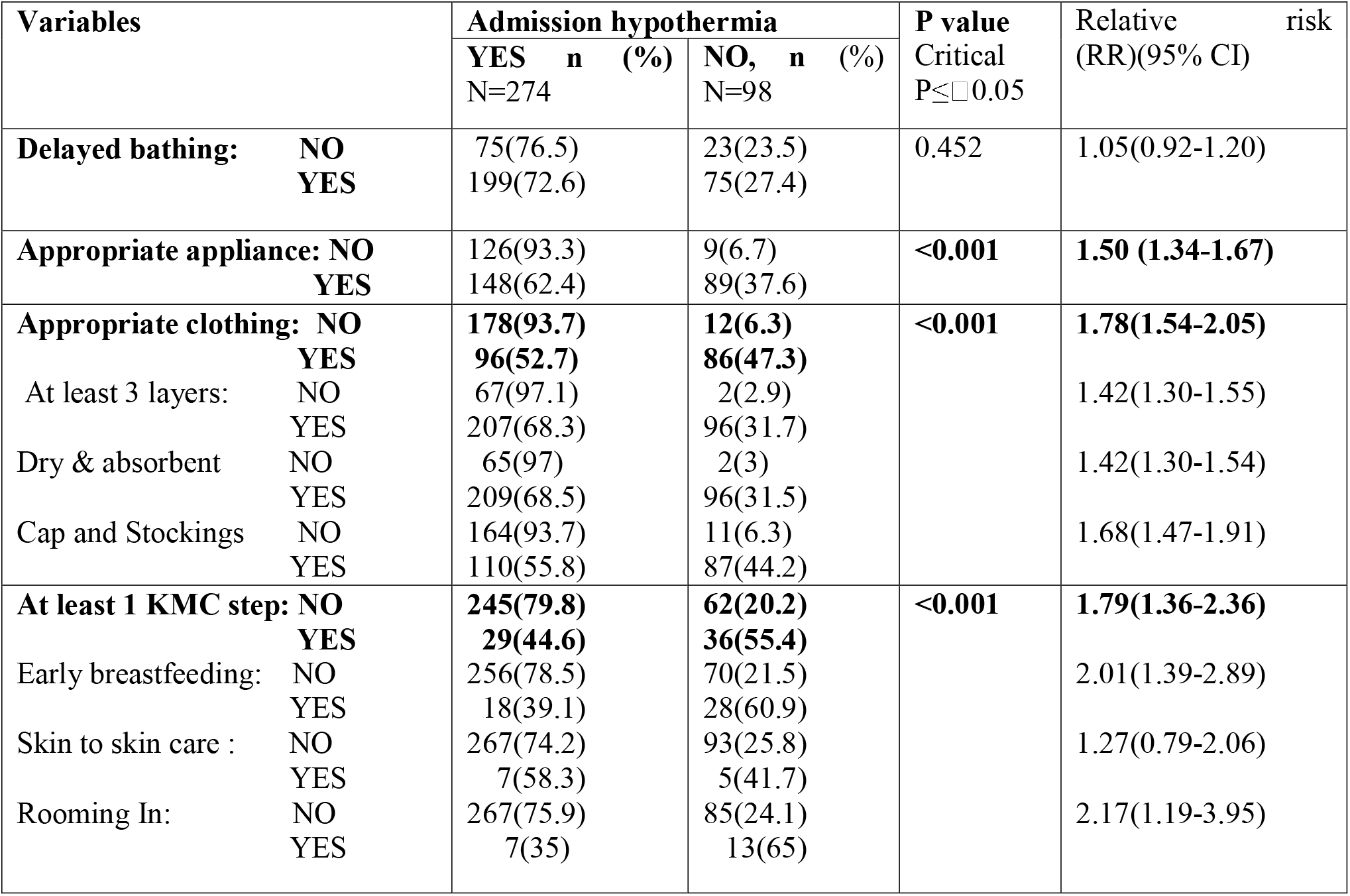
Warm Chain Steps associated with admission hypothermia among neonates admitted at MTRH, Kenya, 2016 [N_D_=_D_372]

### Association between hypothermia, adherence to the warm chain and day-one mortality

Admission hypothermia (RR=17.17; 95% CI: 2.40-122.71), sub optimal adherence to the warm chain by the first hour RR (4.06; 95% CI: 0.5-28.34) and hypothermia recurrence within the initial 24 hours of admission RR (5.23; 95% CI: 1.93-14.19) were significantly associated with day 1 mortality (p<0.001). Admission hypothermia increased the risk of death seventeen times with hypothermia being noted among 48 (98%) of the neonates who died on day one. Hypothermia recurrence and suboptimal adherence to the warm chain by the 1^st^ hour conferred a five -fold and four-fold increased risk of day-one mortality, respectively (Table 5).

**Table 5:** Association between neonatal hypothermia, adherence to the warm chain and day 1 mortality among newborns admitted at MTRH, Kenya in 2016 [N_D_=_D_372]

| Variables | Day one mortality |  | P value | Relative risk (RR)<br>(95% CI) |
| --- | --- | --- | --- | --- |
|  | YES n (%)<br>N=49 | NO. n (%)<br>N=323 |  |  |
| Admission hypothermia<br>YES<br>NO | 48(17.5)<br>1(1) | 226(82.5)<br>97(99) | <0.001 | 17.17(2.40-122.71) |
| Hypothermia recurrence<br>YES<br>NO | 209 (82.3)<br>114 (96.6) | 45 (17.7)<br>4 (3.4) | <0.001 | 5.23 (1.93-14.19) |
| Sub Optimal adherence to warm chain<br>YES<br>NO | 48 (14)<br>1 (3.4) | 295 (86)<br>28 (96.6) | <0.001 | 4.06(0.5-28.34) |

### Association between hypothermia, adherence to the warm chain and day one survival

Absence of hypothermia at admission (AOR=20.907; 95% CI: 2.152 -153.620) and recurrent episodes of hypothermia (AOR=6.136; AOR=2.152 − 17.496) significantly (p<0.001) increased the chances of day one survival, with a twentyfold and six fold increased odds of survival respectively. Optimal adherence to the warm chain by the first hour, significantly improved the chances of day one survival four times albeit without statistical significance (Table 6).

**Table 6:** Association between neonatal hypothermia, adherence to the warm chain and day one survival among newborns admitted at MTRH, Kenya, 2016 [N_D_=_D_372]

|  | Day 1 Survival |  | p-value | Adjusted Odds Ratio<br>(AOR) |
| --- | --- | --- | --- | --- |
|  | YES<br>n=323(%) | NO<br>N=49(%) |  |  |
| Admission hypothermia<br>NO<br>YES | 97 (99)<br>226 (82.5) | 1 (1)<br>48 (17.5) | <0.001 | 20.907 (2.152 -153.620) |
| Hypothermia recurrence<br>NO<br>YES | 114 (96.6)<br>209 (82.3) | 4 (3.4)<br>45 (17.7) | <0.001 | 6.136 (2.152 – 17.496) |
| Adherence to warm chain<br>YES<br>NO | 28 (96.6)<br>295 (86) | 1 (3.4)<br>48 (14) | <0.001 | 4.556 (0.606 – 34.270) |

## DISCUSSION

### Neonatal hypothermia prevalence

Hypothermia was prevalent among neonates admitted at MTRH with 3 out of every 4 newborns having hypothermia at admission. The high proportion of hypothermia in our study could be attributed to the frequency of the associated factors among our participants including prematurity and suboptimal adherence to the warm chain. Our prevalence results matched those from Malaysia (64.8%) [10]Nigerian (72.4%) [11] and Ethiopian 69.8% [12] NICUs. Similar challenges to provision of thermal care are expected among these hospital studies due to climatic, economic, and technological semblance hence the parallel. Lower prevalence rates of 32%,[13] and 51% [14] were recorded in Brazil, while higher prevalence rates were noted in the Netherlands (93%) [15], Nepal (92.3 %) [16], Zimbabwe (85%) [17] and Uganda (83%) [18]. In a local study done in a Kenyan county-referral hospital, a higher prevalence (87%) was reported [19]. The variance in hypothermia rates in these studies could be attributed to differences in temperature measuring sites, timing of thermometry, technological, economic, cultural, and ecological disparities between the study areas and the uniqueness in demographic profiles of the participants.

### Adherence to the WHO newborn thermal care guidelines

There was sub optimal adherence the warm chain at MTRH with less than a tenth of the participants in our study had the warm chain optimally adhered to by the first hour of admission. This is parallel to findings in Malaysia where none of the NICUs evaluated practiced a complete thermal care bundle [10]. Similarly, in Nepal only 10.7% of the neonates in a community study got optimum care [20]. Reviews in Africa also cite negative and sub-standard thermal protection among the factors sustaining the epidemic of hypothermia in the region [7,21]. Provision of appropriate clothing was fairly achieved among our study population credited to the fact that the MTRH newborn unit stocked warm absorbent linen. Very few of the preterm infants weighing less than 1700 grams in our study accessed incubator care as recommended by the WHO due to an overall strain on the seven functional incubators available at the unit during the study period. Incubator sharing was rampant among the preterm infants which confirms observations that use of appropriate thermal care devices is not an optimized practice in Africa [21] unlike in European surveys which map a more technologically advanced economy[22].

### Factors associated with admission hypothermia

Gestational maturity status was significantly associated with hypothermia which concurs with findings in the Netherlands where neonates with a gestational age of less than 32 weeks had 93% hypothermia rates [23]. In Nigeria, 82.5%, (RR=1.51,95% CI: 1.21-1.89) of the hypothermic newborns were preterm [11] with proportions similar to Ethiopian study (OR=4.81;95% CI: 2.67-8.64, p-value =0.001) and an East African meta-analysis (AOR=4.01; 95% CI: 3.02-5.00) [24]. The risk of hypothermia among preterm neonates is innate in their physiology that limits their capacity for thermogenesis as compared to the term infant. Birth weight less than 2500 grams significantly increased the chances of admitted newborns being hypothermic (p<0.001).This matched findings in Nepal (AOR = 4.32; 95% CI: 3.13-5.00) where babies weighing less than 2000g had a four-fold increased risk of hypothermia [25]. In the university of Iowa hospitals and clinics in the United States of America, a high rate of hypothermia (79%) was noted among very low birth weight infants [26]; whereas in Nigeria 93.3% of hypothermic babies had lower than normal weight respectively [11,27]. These findings were replicated in an Ethiopian study where weight was associated with increased odds of hypothermia (AOR=1.33, 95% CI:0.75-2.36) [28]. Sub optimal application of the warm chain was significantly associated with hypothermia. Comparatively, inadequate clothing among hypothermic neonates [21] doubled odds of hypothermia among neonates in Malawi who did not wear caps [29]. Increased odds of hypothermia were further noted among neonates who lacked skin-to-skin care (AOR=2.8 95% CI: 1.46-5.66) [12] as well as those with delayed breastfeeding (AOR=7.58, 95% CI: 3.61-15.91) [12].Similarities in setting and the congruence in challenges faced among the hospital studies could explain the parity.

### Factors associated with day one mortality and survival

There was a marked increase in the risk of day one mortality among newborns who were hypothermic at admission with increased odds of survival among non-hypothermic neonates. Similarly, a higher CFR is noted among hypothermic babies in Nigeria (37.6%) [11] vs. South Africa (16.7%). The relationship of hypothermia and death among neonates is inherent in its pathophysiologic mechanisms which involves compromises in the neonatal biological systems resulting in a cascade of vicious and deleterious events that are fatal.

## CONCLUSIONS AND RECOMMENDATIONS

There was sub optimal adherence to the WHO thermal care guidelines at the MTRH newborn unit with less than 10% achieving optimal adherence to warm chain steps feasible from point of admission by first hour of admission. Prematurity and adherence to the warm chain were significantly associated with admission hypothermia. There was a seventeen-fold increase in the risk of death among neonates who had hypothermia at admission.

There is need to optimize the application of the warm chain at MTRH through quality improvement strategies addressed to the weak links identified by this study. An anticipatory approach to thermal care and priority triage of the preterm neonate admitted at the MTRH newborn unit should be adopted.

## Data Availability

The data from this study will be made publicly available.

## ACKNOWLEDGEMENTS

The authors would like to thank the mothers and the neonates who participated in this study. Secondly, this study could not have happened without the approval and support of Moi Teaching and Referral Hospital’s administration and pediatrics department.

